# Agreement between arterial and capillary pH, pCO2 and lactate in patients in the emergency department

**DOI:** 10.1101/2020.06.11.20128223

**Authors:** V. Collot, S. Malinverni, E. Schweitzer, J. Haltout, P. Mols, M. Bartiaux

## Abstract

**Study objective:** The primary objective of the study was a quantitative analysis to assess the mean difference and 95% confidence interval of the difference between capillary and arterial blood gas analyses for pH, pCO2 and lactate. Secondary objective was to measure the sensitivity and specificity of capillary samples to detect altered pH, hypercarbia and lactic acidosis.

**Methods:** Adults admitted to the ED for whom the treating physician deemed necessary an arterial blood gas analysis (BGA) were screened for inclusion. Simultaneous arterial and capillary samples were drawn for BGA. Agreement between the two methods for pH, pCO2 and lactate were studied with Bland-Altman bias plot analysis. Sensitivity, specificity, positive and negative predictive value as well as AUC were calculated for the ability of capillary samples to detect pH values outside normal ranges, hypercarbia and hyperlactatemia.

**Results:** 197 paired analyses were included in the study. Mean difference for pH, between arterial and capillary BGA was 0.0095, 95% limits of agreement were -0.048 to 0.067. For pCO2, mean difference was -0.3 mmHg, 95% limits of agreement were -8.5 to 7.9 mmHg. Lactate mean difference was -0.93 mmol/L, 95% limits of agreement were -2.7 to 0.8 mmol/L. At a threshold of 7.34 for capillary pH had 98% sensitivity and 97% specificity to detect acidemia; at 45.9 mmHg capillary pCO2 had 89% sensitivity and 96% specificity to detect hypercarbia. Finally at a threshold of 3.5 mmol/L capillary lactate had 66% sensitivity to detect lactic acidosis.

**Conclusion:** Capillary measures of pH, pCO2 and lactate can’t replace arterial measurements although there is high concordance between the two methods for pH and pCO2 and moderate concordance for lactate. Capillary blood gas analysis had good accuracy when used as a screening tool to detect altered pH and hypercarbia but insufficient sensitivity and specificity when screening for lactic acidosis.

## Introduction

### Background

Arterial blood gas analysis (BGA) is universally used in emergency departments (ED) to assess for acid-base abnormalities, oxygenation and ventilation status in patients presenting with suspected severe illnesses. Arterial BGA involve an arterial puncture or the placement of an indwelling arterial catheter. Both techniques are painful^1^, can be distressing for the patients are associated with potential complications in a significant proportion of cases, such as hematomas, arterial lesions, ischemia, nervous lesions, algoneurodystrophya or infections^2,3፧^.

Alternatives to arterial BGA of proven efficacy exist. Continuous pulse oximetry is ubiquitously used for monitoring oxygenation and has been proven both accurate and precise when compared to SaO2. Venous BGA, with or without mathematical arterialization of the results, has been shown to be both moderately accurate and highly precise in terms of prediction of arterial pH^4–8^. Venous pCO2 values with a threshold of 45 mmHg can be used as a highly sensible screening test for hypercarbia in COPD patients^9^፧ but venous pCO2 values show only moderate accuracy on predicting arterial pCO2^4,5,7^ and cannot be used to replace arterial pCO2 values^8,10^. The agreement between venous and arterial lactate is sufficient only to rule out hyperlactatemia in septic patient when absent in venous blood^11^. Altogether these results suggest that VBG could be used as a screening tool in emergency care when coupled with clinical reasoning^12^ but better alternatives to ABG than VBG should be investigated.

Capillary blood sampling for BGA is frequent and widely adopted for the assessment of the acid-base status and pCO2 values in pediatric patients worldwide^13–16^. This technique is associated with extremely rare complications^17^, a reduced discomfort for the patient^18^ and an easier sampling technique requiring less technically qualified personnel. In the adult population, capillary BGA, is not as widely disseminated as for the pediatric population. In adults BGA on capillary blood was studied both with normal and arterialized samples. Results from arterialized samples, obtained by warming the skin or applying a vasodilator substance, showed a good agreement between the two techniques in terms of pH and pCO2^18–20^. Nevertheless this technique is time consuming and work-intensive preventing its use in ED beyond research purposes. Moreover a recent study from an ICU population seem to show that similar results could be obtained without prior arterialization of the capillary bed^21^. A metanalysis showed encouraging results from capillary BGA but given the extremely heterogeneous population composed partially of perfectly healthy subjects these results cannot be extrapolated to emergency care. Moreover studies conducted in the ED to specifically study capillary BGA only reported correlation coefficients without studying mean difference and 95% limits of agreement, therefore answering incompletely to the question on whether capillary BGA could replace arterial BGA or could be used as a reliable alternative^22^.

The primary objective of the study was to assess the agreement between capillary BGA and the reference gold standard technique arterial BGA in terms of pH, pCO2 and lactate through Bland-Altman agreement plot technique^23^. Secondary outcome of our study was to identify and describe the performance of capillary BGA to detect altered pH, hypercarbia and lactic acidosis at different thresholds.

## Methods

We conducted a prospective monocentric study on a convenience sample of patients admitted to the ED from December 2017 to April 2018. Patients aged 18-99 presenting to the ED for whom the treating clinician deemed necessary an arterial blood gas analysis (BGA) were screened for inclusion in the study if not pregnant. Screening for inclusion was performed according to study personnel availability and overall workload in the ED. We calculated a sample size of 200 patients based on previous study^21^ reporting a standard deviation of differences for pH of 0.0338. We based our sample calculation on the precision of the estimates and aimed for a standard error of the 95% limit of agreement of approximatively +/-0.01.

Simultaneous arterial and capillary samples were drawn for BGA. Arterial BGA were sampled by direct arterial radial or femoral puncture using a pre-heparinized syringe (safe-PICO aspirator®, 1,7ml, Radiometer® and a 23G needle (BD Microlance® 3, Becton Dickinson®). Capillary samples were obtained at patients finger using a contact-activated lancet (BD Microtainer®, Becton dickinson®) and collected through capillary tubes (safeCLINITUBES® 70µl, Radiometer®). Samples were analyzed with the same Point of care BGA analyzer (ABL90 FLEX® Radiometer®). Analysis of samples was not blinded, clinical information, index test results and reference standard results were available to the assessors of the reference standard.

Agreement between the two methods for pH, pCO2 and lactate were studied with Bland-Altman bias plot analysis. Upper and lower 95% limits of agreement were calculated as the mean difference +/-1.96 standard deviation of the difference. Sensitivity, specificity, positive and negative predictive values as wells as AUC were calculated for the ability of capillary samples to detect pH values outside normal ranges, hypercarbia and hyperlactatemia. We considered acidemia as a pH below 7.35, alkalemia as a pH above 7.45, hypercarbia as a pCO2 above 45 mmHg and hyperlactatemia as a lactate above 2 mmol/L.

Reported analytical error for pH following an internal quality control showed a standard deviation of 0.0016 for repeated measures at 7.17. Reported analytical error for pCO2 measurements are associated with a standard deviation of 0.228 when pCO2 was 65.9 mmHg. Reported analytical error for lactate was associated with a standard deviation of 0.52 when lactate was at 3.55 mmol/L.

We considered that a capillary measure could replace an arterial measure if the double of the standard deviation of the bias of pH, pCO2 or lactate would be inferior to the reference analytical error (bias + 1.65SD) of the laboratory for the parameter at study.

A post-hoc subgroup analysis to identify variables accounting for variability in diagnostic accuracy was performed.

Missing data were handled as missing and no imputation was performed.

The study was approved by the Ethical Committee of Saint Pierre (B076201630203) and informed consent was obtained from all participants. Study was supported by an unconditional grant from Association pour l’aide à la recherche médicale André Vésale.

## Results

During the study period 1298 blood gas analysis on 1170 patients were performed in our ER. Between the 18^th^ of December 2017 and April 2018, 242 patients underwent screening for participation generating 285 pairs of arterial and capillary BGA. 41 declined consent and one patient was excluded as discovered pregnant after initial inclusion. In 13 cases the arterial sample was inappropriate or unavailable for analysis: in three patients no arterial blood could be obtained and in ten cases blood was a mixture of venous and arterial. In 33 cases the capillary sample was inappropriate for analysis: in 15 cases blood drawn in the capillary tube was insufficient for analysis, in six cases capillary blood could not be obtained from the patient, in five cases air bubbles were present in the capillary tube and in five other patients unspecified problems hampered the analysis. After screening for exclusion criteria a total of 197 paired BGA, accounting for 167 patients were included in our study (Figure 1). The mean age of the 167 study participants was 57.9 years (+/-16.3) and 126 participants (63.9%) were men. Baseline characteristics, hemodynamic variables and clinical diagnoses in ED are reported in Table 1. Acidemia, defined as a pH below 7.35, was present in 47 arterial samples (23.9%) while alkalemia, defined as a pH above 7.45, was present in 41 (20.9%). Hypercarbia, defined as a pCO2 above 45 mmHg, was present in 66 arterial samples (33.5%) and elevated lactate, defined as a lactate above 2 mmol/L, was found in 51 (25.9%). Median delay between arterial and capillary was one minute (range, 0 to 4).

**Figure 1.**
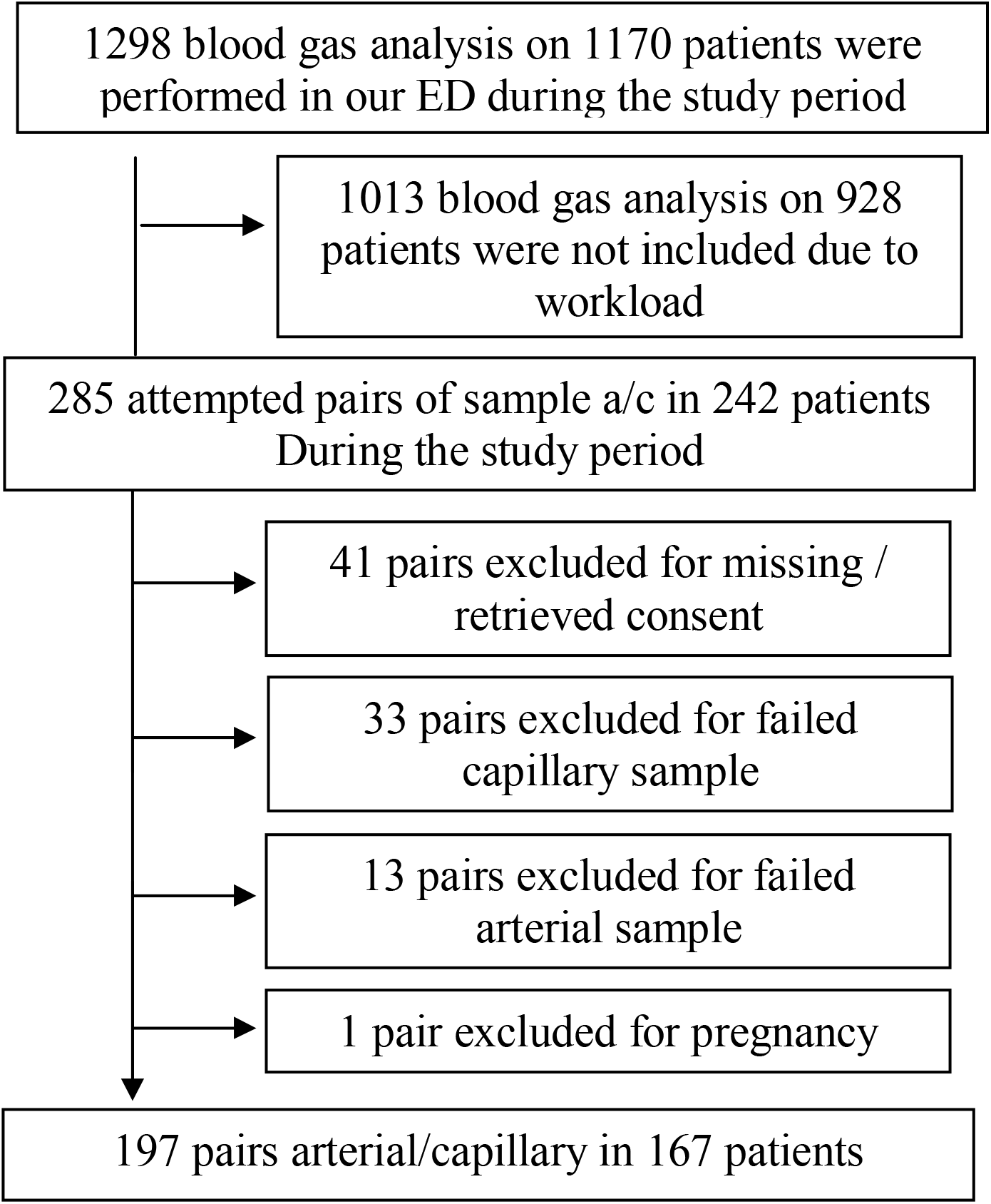
Inclusion flowchart

**Table 1.** Demographic and clinical characteristics of patients at study inclusion

|  |  |
| --- | --- |
| *Number of observations* | 197 |
| Age, mean (SD), y | 57.9 (16.3) |
| Male, No. (%) | 126 (63.9) |
| BMI, median (SD) | 24.7 (20.8 – 28.5) |
| **Suspected pathology at sampling, No. (%)** |  |
| COPD | 52 (26.4) |
| Asthma | 7 (3.6) |
| Pneumonia | 16 (8.2) |
| Pulmonary embolism | 4 (2) |
| Other respiratory failure | 22 (11.2) |
| Carbon monoxide poisoning | 3 (1.5) |
| Diabetic ketoacidosis | 16 (8.1) |
| Epilepsia | 12 (6.1) |
| Sepsis | 12 (6.1) |
| Shock | 6 (3.1) |
| ROSC | 3 (1.5) |
| Cardiac failure | 15 (7.6) |
| CVA | 1 (0.5) |
| Other | 28 (14.2) |
| **Capillary blood sampling attempts, No (%)** |  |
| One attempt | 170 (92.9) |
| Two attempts | 12 (6.6) |
| Three or more attempts | 1 (0.5) |
| **Arterial blood sampling attempts No. (%)** |  |
| One attempt | 165 (90.7) |
| Two attempts | 13 (7.1) |
| Three or more attempts | 4 (2.2) |
| Mean arterial pressure (SD), mmHg | 96.0 (21.1) |
| Delay arterial - capillary (IQR), min | 1 (1 - 2) |

Abbreviations: BMI, body mass index; COPD, chronic obstructive pulmonary disease; CVA, cardiocascular accident; IQR, interquartile range; ROSC, return of spontaneous circulation SD, standard deviation

Median pH was 7.41 (IQR 7.36 – 7.44). Mean difference (i.e. bias) for pH (arterial – capillary) was 0.01 with 95% limits of agreement going from -0.048 to 0.067 (Figure 2A). Concordance correlation coefficient between arterial and capillary pH was 0.943 (95% CI, 0.928 – 0.959). Percentage error was 0.79%.

Median pCO2 was 40.8 mmHg (IQR 34.7 – 47.3). The mean pCO2 bias (arterial – capillary) between paired methods was -0.3 mmHg with 95% limits of agreement going from -8.5 to 7.9 mmHg (Figure 2B). The concordance correlation coefficient between arterial and capillary pCO2 was 0.963 (95% CI, 0.952 -0.974). Percentage error was 18.9%.

Median lactate was 1.2 mmol/L (IQR 0.8 – 2.1).The mean lactate bias (arterial – capillary) between paired methods was -0.93 mmol/L with 95% limits of agreement going from -2.7 to 0.84 mmol/L (Figure 2C). The concordance correlation coefficient between arterial and capillary lactate was 0.824 (95% CI, 0.783 – 0.865). Percentage error was 78.8%.

The area under the receiver operating characteristic (ROC) curve constructed to assess the ability of capillary blood gas analysis to detect for arterial acidemia was 0.99 (95% CI, 0.99 to 1) (Figure 3). Using a cutoff point of 7.34 for capillary pH to detect acidosis sensitivity was 98% and specificity was 97%. The positive predictive value of a capillary pH below 7.34 to detect acidemia was 92% while the negative predictive value was 99%.

When studying our secondary outcomes, the area under the ROC curve constructed to assess the ability of capillary blood gas analysis to detect for arterial alkalemia was 0.94 (95% CI, 0.90 to 0.97). With a cutoff point of 7.447 for capillary pH to detect arterial alkalosis sensitivity was 71% and specificity 94%. The positive predictive value of a capillary pH above 7.447 to detect alkalemia was of 79% while the negative predictive value was 91%.

The area under the ROC curve constructed to assess the ability of capillary blood gas analysis to predict for arterial hypercarbia was 0.97 (95% CI, 0.95 – 0.99). Using a cutoff of 45.9 mmHg for capillary pCO2 to detect hypercarbia sensitivity was 89% while specificity was 96%. The positive predictive value of a capillary pCO2 above 45.9 mmHg to detect hypercarbia was of 92% while the negative predictive value was 94%.

Finally, the area under the ROC curve constructed to assess the ability of capillary blood gas analysis to detect arterial hyperlactatemia was 0.90 (95% CI 0.85 - 0.95). A threshold of 3.5 mmol/L for capillary lactate was associated with a sensitivity of 66% and a specificity of 93% for detecting hyperlactatemia. The positive predictive value of a capillary lactate above 3.5 mmol/L was 76% while the negative predictive value of 89%.

Sensitivity for detecting acidemia was 100% when using a pH cut-off value of 7.36 (Table 2). This cut-off value was associated with a specificity of 93% and a positive predictive value of 0.82.

**Table 2.** Cutoff Levels, Sensitivity and Specificity for capillary BGA in detecting altered pH, pCO2 or lactate

| Value | Cutoff Level | Sensitivity % (95% CI) | Specificity % (95% CI) | Patients, No. (%) |  |  |  | PPV % (95% CI) | NPV % (95% CI) |
| --- | --- | --- | --- | --- | --- | --- | --- | --- | --- |
|  |  |  |  | True Positive | False Negative | False Positive | True Negative |  |  |
| Arterial pH [acidosis] | 7.361 | 100<br>(92.4 - 100) | 93.3<br>(88.1 - 96.8) | 47<br>(23.9) | 0<br>(0) | 10<br>(5.1) | 140<br>(71.1) | 82.5<br>(72.1 - 89.5) | 100 |
|  | 7.34 | 97.9<br>(88.7 - 100) | 97.3<br>(93.3 - 99.3) | 46<br>(23.7) | 1<br>(0.5) | 4<br>(2.0) | 146<br>(74.5) | 92<br>(81.4 - 96.8) | 99.3<br>(95.5 - 99.9) |
|  | 7.288 | 53.1<br>(38.1 - 67.9) | 100<br>(97.6 - 100) | 25<br>(12.7) | 22<br>(11.2) | 0<br>(0) | 150<br>(76.6) | 100 | 87.2<br>(83.4 - 90.2) |
| Arterial pH [alkalosis] | 7.4 | 100<br>(91.4 - 100) | 62.2<br>(54.1 - 69.8) | 41<br>(20.8) | 0<br>(0) | 59<br>(30) | 97<br>(49.2) | 41.0<br>(36.2 - 45.9) | 100 |
|  | 7.447 | 65.9<br>(49.4 - 79.9) | 95.5<br>(91.0 - 98.2) | 27<br>(13.1) | 14<br>(7.1) | 7<br>(3.5) | 149<br>(75.6) | 79.4<br>(64.4 - 89.2) | 91.4<br>(87.4 - 94.2) |
|  | 7.471 | 39.0<br>(24.2 - 55.5) | 100<br>(97.7 - 100) | 16<br>(8.1) | 25<br>(12.7) | 0<br>(0) | 156<br>(79.2) | 100 | 86.2<br>(83.0 - 88.9) |
| pCO2 (mmhg) [hypercapnia] | 35.8 | 100<br>(94.6 - 100) | 42.0<br>(33.4 - 50.9) | 66<br>(33.5) | 0<br>(0) | 76<br>(38.6) | 55<br>(27.9) | 46.5<br>(42.9 - 50.1) | 100 |
|  | 45.9 | 89.4<br>(79.4 - 95.6) | 96.2<br>(91.3 - 98.8) | 59<br>(29.9) | 7<br>(3.6) | 5<br>(2.5) | 126<br>(63) | 92.2<br>(83.3 - 96.6) | 94.7<br>(89.9 - 97.3) |
|  | 56.5 | 51.5<br>(38.9 - 64.0) | 100<br>(97.2 - 100) | 34<br>(17.3) | 32<br>(16.2) | 0<br>(0) | 131<br>(66.5) | 100 | 80.4<br>(76.2 - 84.0) |
| Lactate (mmol/L) [hyperlactatemia] | 1.3 | 100<br>(92.5 - 100) | 23.0<br>(16.2 - 31.0) | 47<br>(25.8) | 0<br>(0) | 104<br>(57.1) | 31<br>(17.3) | 31.1<br>(29.2 - 33.1) | 100 |
|  | 3.5 | 66.0<br>(50.7 - 79.1) | 92.6<br>(86.8 - 96.4) | 31<br>(17.3) | 16<br>(8.8) | 10<br>(5.5) | 125<br>(68.7) | 75.6<br>(62.3 - 85.4) | 88.7<br>(84.0 - 92.1) |
|  | 4.4 | 46.8<br>(32.1 - 61.9) | 100<br>(97.3 - 100) | 22<br>(12.1) | 25<br>(13.7) | 0<br>(0) | 135<br>(74.2) | 100 | 84.4<br>(80.5 - 87.6) |

Abbreviations: PPV, Positive Predictive Value; NPV, Negative Predictive Value

Sensitivity for detecting hypercarbia was 100% when using a pCO2 cut-off value of 35.8 mmHg. This cut-off value was associated with a specificity of 42% and a positive predictive value of 0.47.

For lactate, sensitivity for detecting hyperlactatemia was 100% when using a cut-off value of 1.3 mmol/L. In this case specificity was 23% and the positive predictive value was 0.31.

A subgroup analysis according to delays between samples, presence of hypotension and suspected pathology did not yield any evident difference between subgroups (Table 3).

**Table 3.**
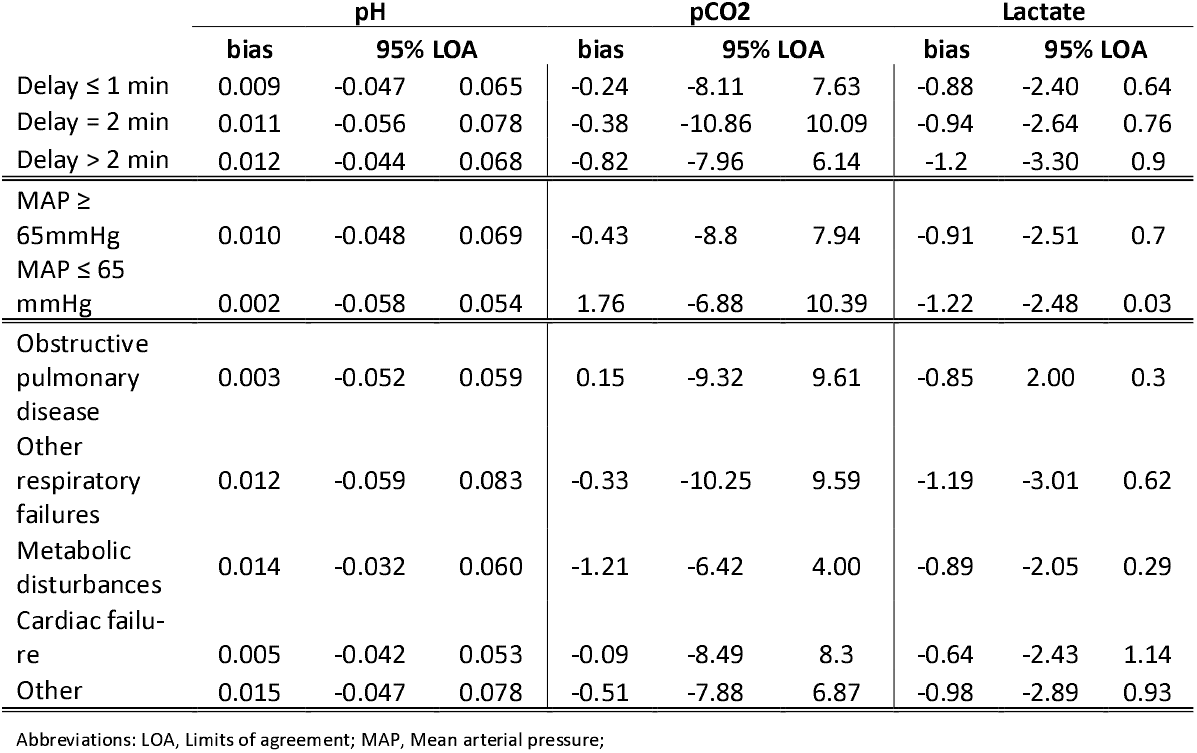
Subgroup analysis according to delays in sampling and suspected diagnosis

Data supporting the study are available at 10.6084/m9.figshare.12464888.

## Discussion

The result of this study indicate that capillary blood sampling to assess pH and pCO2 show high accuracy and moderate precision when compared to the gold standard technique of arterial blood sampling. Accuracy and precision in terms of predicting arterial lactate from capillary values were lower in our sample.

In our study the double of the standard deviation of the bias of pH, pCO2 and lactate was superior to the reference analytical error (bias + 1.65SD) of the laboratory for the parameter at study. Capillary sampling cannot therefore alone simply replace arterial sampling for blood gas analysis in an ED.

Nevertheless according to ROC curves analysis a capillary pH is highly predictive of arterial acidemia. A capillary pH below 7.36 yielded a sensitivity of 100% and a specificity of 93% for detecting acidemia meaning a negative predictive value of 100%. Capillary blood gas analysis with a threshold of a pH=7.36 could be used for patients in respiratory distress with suspected respiratory acidosis as a triage tool to screen for patients needing further assessment through arterial blood gas analysis. Patient with a capillary pH below 7.36 would be further assessed through an arterial blood gas analysis and quickly treated with non-invasive ventilation if coherent with patient history and clinical exam. This could safely avoid useless, painful and potentially harmful arterial sampling in a substantial proportion of patients presenting in the ED for a respiratory condition. In our sample, within 59 patients presenting for an obstructive pulmonary condition, capillary blood gas analysis with a threshold of 7.36, would have avoided 37 (62.7%) arterial blood sampling assuming the aim of the analysis was to detect respiratory acidosis. Using the same threshold out of 16 patients with decompensated diabetes suspected for diabetic ketoacidosis capillary blood sampling would have avoided 15 (93.8%) arterial sampling assuming that the aim of the analysis was to detect arterial acidemia only.

A recent meta-analysis of studies comparing venous BGA to arterial BGA showed a mean difference for pH between the two techniques of 0.033 and limits of agreement ranging from -0.023 to 0.090^8^, in line with what reported from other studies focused on the ED ^4,24,25^. Our study shows a lower mean difference of 0.01 and similar limits of agreement going from -0.048 to 0.067 when comparing arterial BGA to capillary BGA suggesting that the latter might be a better alternative than venous BGA. When comparing pCO2 values drawn in the ED from venous BGA to values obtained from arterial BGA a recent review article showed a mean difference for pCO2 between the two techniques of 5.7 mmHg with limits of agreement falling outside ± 10 mmHg in most of the cases^24^. Our study again show a mean difference of -0.3 mmHg and limits of agreement going from -8.5 to 7.9 mmHg when comparing pCO2 from arterial BGA to pCO2 from capillary BGA suggesting again that the latter might be a more accurate alternative than venous BGA.

Our study shows encouraging results in terms of accurate detection of arterial hypercarbia through capillary BGA with an AUC of 0.97. When using a threshold of 45.9 mmHg, capillary BGA had a positive predictive value of 92% and with a negative predictive value of 94% for detecting hypercarbia (Table 2).

According to study results capillary lactate levels are only moderately accurate and precise in their prediction of arterial lactate levels. Capillary lactate levels tend to be systematically higher than arterial levels and with a percentage error of almost 80% which is largely above any cutoff level commonly considered as acceptable when using a precise reference method. Lactate measurement by CBG did not have adequate agreement with ABG for clinical use.

Altogether these results show that capillary blood gas analysis could be reliably used as an almost painless screening tool for acidemia, alkalemia and hypercarbia in patients suspected for alteration of both pH and pCO2 in the ED. This could avoid a substantial proportion of arterial blood gas analysis reducing not only the number of painful exams to which a patient is exposed in the ED but could also streamline the patient flow within the ED. This would be achieved by replacing cumbersome and time consuming exams requiring skilled personnel such as for arterial blood gas analysis with screening exams easier to perform and less skill-intensive.

Overall our results support that capillary BGA can be safely integrated in the routine practice of an ED as a screening tool to identify patients with altered pH, hypercarbia and elevated lactate level and that this could safely reduce the number of unnecessary arterial BGA. These results urgently question some well rooted routine procedures as being in opposition with the first principle of Hippocratic medicine (first do not harm) since less painful, safe and precise alternatives exist for the initial screening of patients in the ED presenting with suspected severe conditions.

Cost associated with capillary sampling may differ between regions and may be context specific. In our context capillary BGA material and analysis costs ware only slightly higher than for arterial BGA. Moreover no additional equipment was needed except for the capillary tubes themselves in our study. Given the gain associated with capillary BGA both in terms of workload and workforce cost or the need of additional equipment should not restrain capillary BGA implementation in emergency departments.

Our study has both strengths and limitations. One limitation of our study is its monocentric design. Our monocentric design limits the ability the possibility to generalize our results to other emergency departments in other settings. Second limitation is having used a convenience sample of ideally 200 patients. Patients could not be systematically screened and inclusion was erratic and according to workload. This led to the exclusion of a majority of patients consulting during the study period and might have introduced a bias in recruitment. The main strengths of our study are its pragmatic design with wide study population both in terms of size and variety of pathologies together with his rigorous reporting of results well beyond correlation analysis. Further studies should replicate our findings in different settings and test the implementation of capillary blood gas analysis for screening purposes.

## Conclusion

Capillary samples for blood gas analysis have a high concordance for pH and pCO2 and moderate concordance for lactate when compared to arterial sampling. However precision is insufficient and capillary cannot replace arterial sampling. Capillary blood gas analysis had good sensitivity and specificity when used as a screening tool to detect altered pH and hypercarbia but insufficient sensitivity and specificity when screening for lactic acidosis.

## Data Availability

Data supporting the manuscript will be made available upon request.

https://doi.org/10.6084/m9.figshare.12464888.v1

---

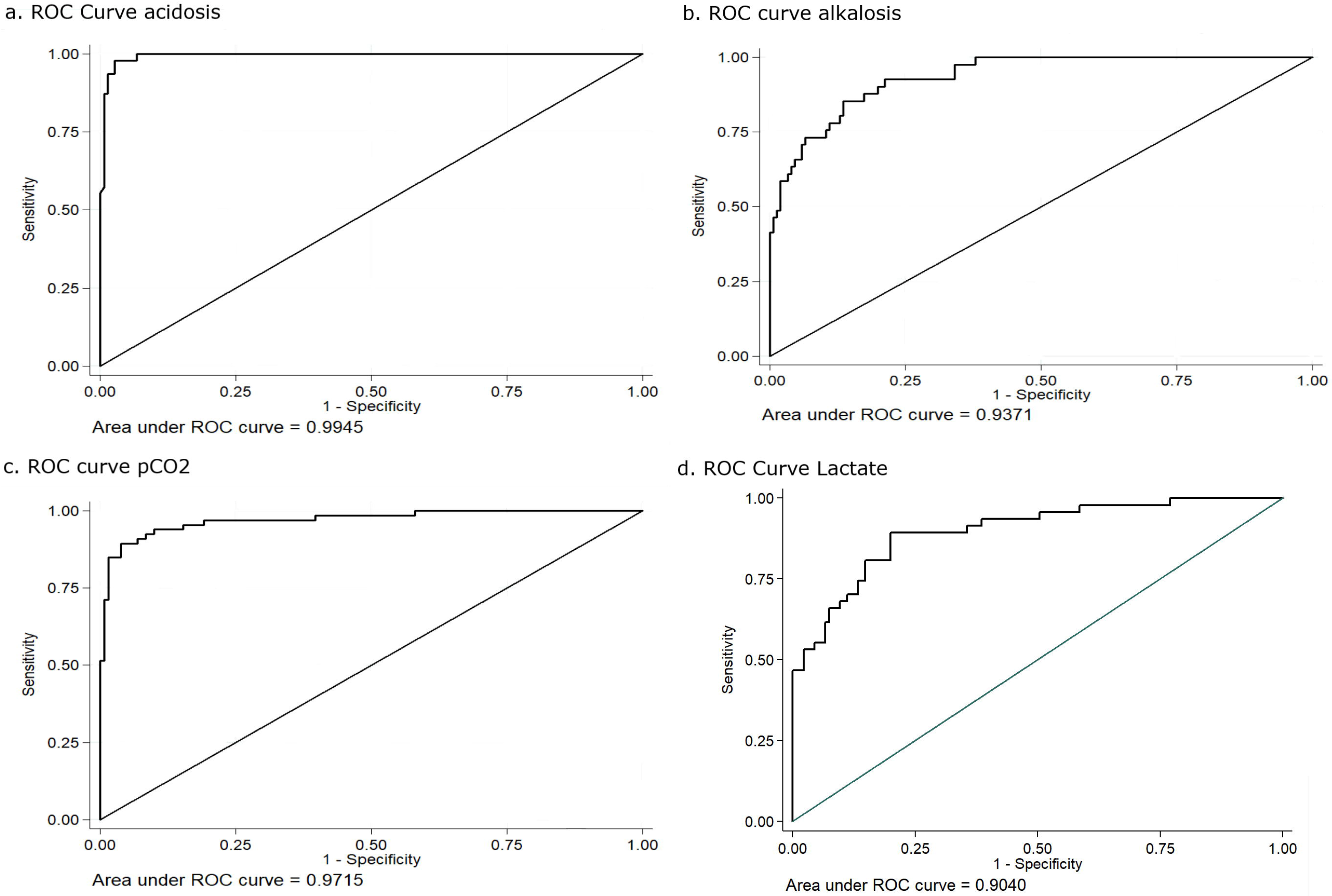

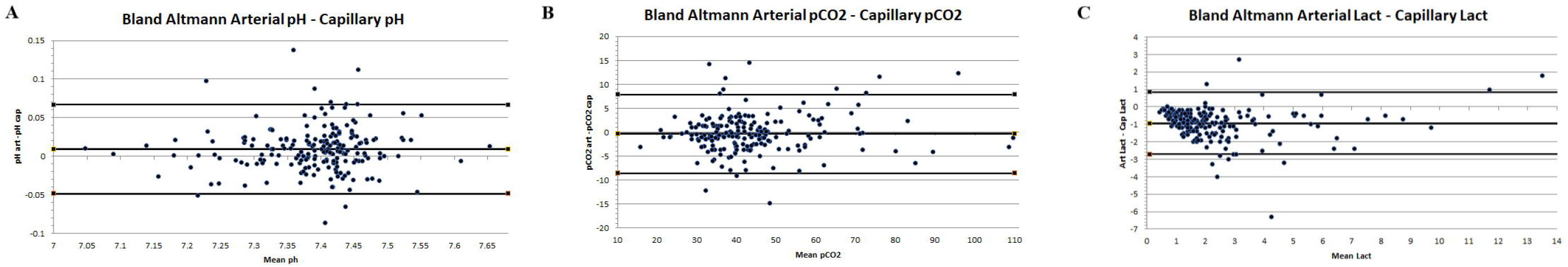

